# Illusory social agents within and beyond voices: a computational linguistics analysis of the experience of psychosis

**DOI:** 10.1101/2021.01.29.21250740

**Authors:** Lisha Shiel, Zsófia Demjén, Vaughan Bell

**Affiliations:** Research Department of Clinical, Educational and Health Psychology, University College London, London, UK; UCL Centre for Applied Linguistics, University College London, London, UK; South London and Maudsley NHS Foundation Trust, London, UK

## Abstract

**Aims:** Psychosis has a strong social component and often involves the experience of being affected by ‘illusory social agents’. However, this experience remains under-characterised, particularly for social agents in delusions and non-vocal hallucinations. One useful approach is a form of computational linguistics called corpus linguistics that studies texts to identify patterns of meaning encoded in both the semantics and linguistic structure of the text.

**Methods:** Twenty people living with psychosis were recruited from community and inpatient services. They participated in open-ended interviews on their experiences of social agents in psychosis and completed a measure of psychotic symptoms. Corpus linguistics analysis was used to identify key phenomenological features of vocal and non-vocal social agents in psychosis.

**Results:** Social agents are i) represented with varying levels of richness in participants’ experiences, ii) are attributed with different kinds of identities including physical characteristics and names, iii) are perceived to have internal states and motivations that are different from those of the participants, and iv) interact with participants in various ways including through communicative speech acts, affecting participants’ bodies and moving through space. These representation were equally rich for agents associated with hallucinated voices and those associated with non-vocal hallucinations and delusions.

**Conclusions:** We show that the experience of illusory social agents is a rich and complex social experience reflecting many aspects of genuine social interaction and is not solely present in auditory hallucinations, but also in delusions and non-vocal hallucinations.

## 1. Introduction

One of the most striking aspects of psychosis is that delusions and hallucinations are strongly social in nature and typically involve the experience of being bothered by, or interacting with, ‘illusory social agents’ (Bell et al., 2017). However, only recently have these experiences been considered to be of phenomenological interest in terms of informing scientific models of psychosis (Alderson-Day et al., 2020; Alderson-Day and Fernyhough, 2016; Bell, 2013; Leudar et al., 1997; Rosen et al., 2016; Wilkinson and Bell, 2016). Notably, however, almost all of these studies have focused on hallucinated voices, rather than delusions or non-auditory experiences in psychosis, and most involve coding broad features from interviews or use survey methodology with pre-selected questions.

Traditionally, fine-grain phenomenological studies of agents in psychosis have used qualitative analysis of open-ended interview transcripts (e.g. Beavan, 2011; Corstens and Longden, 2013) or approaches from phenomenological philosophy (e.g. Humpston and Broome, 2015; Larøi et al., 2010). Both are important but, by design, rely on systematic but subjective analyses that may mean the findings are not reproducible to the same degree as quantitative analyses.

One alternative approach is a form of computational linguistics called corpus linguistics which is the computer-aided study of the systematic patterns in texts to identify patterns of meaning encoded in both the semantics and linguistic structure of the text (McEnery and Hardie, 2011). It involves both statistical and interpretive elements, allowing for analysis of subtle aspects of meaning while potentially being less subjective.

Initial studies have applied this approach to understanding the experience of hallucinated voices in psychosis. Demjén and Semino (2015) initially applied this to the experience of voices as described in a published autobiographical account and later to interviews with 40 voices hearers in an early intervention programme for psychosis (Collins et al., 2020) – reporting how linguistic features represented important features of identity and social interaction with voices that related to voice-related distress. Indeed the social features of voices have been previously identified as being important in driving the distress and disability associated with voice hearing in psychosis (Mawson et al., 2010).

In an attempt to better understand the phenomenology of illusory social agents in psychosis, across voices and other key experiences in psychosis – namely delusions and non-vocal hallucinations – we completed 20 open-ended interviews with patients about their experience of agents in psychosis. We subsequently, analysed the text with corpus linguistics to identify the types of qualities of social agents including how they are perceived to ‘think, behave and interact’.

## 2. Method

### 2.1 Participants

Participants were recruited from a psychosis outpatient service and an acute psychiatric inpatient ward. Participants were invited to participate if they a) were under the age of 65 or over the age of 18; b) were identified by the clinical team as having psychosis; c) were English language speakers; and d) had capacity to consent to the research. The study was approved by the London-Dulwich NHS Research Ethics Committee (Ref: 17/LO/0171).

For this study, 27 participants (13 women; 15 inpatients) were recruited with 20 interviews included in the final analysis. The interviews of seven participants were excluded because after starting the interview they either reported not hearing voices or experiencing delusion-like experiences or were reluctant to discuss these experiences with the interviewer.

### 2.2 Materials and measures

#### 2.2.1 Qualitative interview

This was an open format qualitative interview led by a topic guide that included: experiences of hearing things that others cannot hear, characteristics of the voice/thing that is heard, nature of the relationship between the person and the voice, experience of delusions, exploration of any characters in the delusions and how the participant relates to them, and exploration of links between past experiences and delusions.

#### 2.2.3 Psychotic Symptom Ratings Scales

Auditory hallucinations and delusions were measured using the Psychotic Symptom Ratings Scales: Voices (PSYRATS-V) and Psychotic Symptom Rating Scales: Delusion (PSYRATS-D) scales (Haddock et al., 1999).

#### 2.2.4 Demographic information

Gender, age and ethnicity were recorded.

### 2.3 Procedure

Participants were interviewed in hospital where they had outpatient appointments or in the ward in which they were an inpatient. After discussing the study and agreeing to consent, participants engaged in the open-ended interview which was audio recorded. Interviews were transcribed verbatim and any references to identifiable names, addresses and other personally identifying information were removed to create anonymised transcripts used in the analysis. In the second part, participants provided brief demographic information and completed the PSYRATS with the interviewer. The open-ended interview was completed before the PSYRATS to avoid shaping later responses.

### 2.4 Analysis

Anonymised verbatim transcripts were analysed using a corpus linguistics approach that combines quantitative and qualitative techniques. In particular, the software package *#LancsBox* (version 4) was used to analyse the corpus (Brezina et al., 2018, 2015). The *#Lancsbox* analysis files minus the original transcripts have been made available on the online archive: https://osf.io/4zwq8/

#### 2.4.1 Initial processing

Initial processing of the interview text involved corpus annotation where references to vocal illusory social agents and non-vocal illusory social agents were tagged. References to illusory social agents were identified by both reference to the text and to the PSYRATS assessment that identified psychotic symptoms. Agent tagging was initially completed by the first author and checked by one of the two other authors. Differences were resolved collaboratively by consulting the third author.

The final tagged transcripts were loaded into the *#LancsBox* software package for analysis. A general reference corpus was also uploaded to the software for comparison to the interview corpus. The reference corpus consisted of the Oral History Interviews section of the spoken part of the British National Corpus (BNC Consortium, 2007). This corpus, although historically older, is similar in format to the data in this study in terms of it being transcriptions of semi-structured interviews with people from different demographics, but in a range of contexts.

Keywords are then identified which are statistically significantly more frequent in the clinical corpus when compared to the reference corpus. They allow characterisation of what is typical and specific to a particular dataset (Baker, 2010). We used two statistical measures to generate keywords: (i) Log Likelihood (LL), a test of statistical significance; and (ii) Log Ratio, an effect size statistic, representing the size of the difference between two corpora for each statistically significant item. We used a LL cut-off of 10.83 (p<0.001) and a Log Ratio cut off point of 1.5, which meant that all keywords we considered were at least three times more common in the clinical corpus.

#### 2.4.2 Social agent characterisation

We used the *#LancsBox* keyword in context (‘KWIC’) and collocation (‘GraphColl’) analyses to characterise the illusory social agents described in the clinical interview texts.

##### 2.4.2.1 Types of social agents

We used the keyword in context (‘KWIC’) analysis to concordance (i.e. list each occurrence with the words surrounding it) all references to i) vocal illusory social agents, and ii) non-vocal illusory social agents. This allowed us to determine the range of references (i.e. the number of ‘types’), their frequency, and how they were used.

##### 2.4.2.2 Qualities of social agents

To understand how social agents were described as being experienced and perceived by participants we used collocation analysis using the *#LancsBox* GraphColl function. Collocations are combinations of words that frequently co-occur in a corpus (Brezina et al., 2015) and represent the idea that important aspects of meaning are not contained within individual words but can be found in the characteristic associations of a word (i.e. the company it keeps), including other words and structure with which it frequently co-occurs. We examined adjective collocates that highlighted how social agents are described in the corpus, and verb collocates, which gave a sense of the activities and behaviours that social agents reportedly engaged in. Verb collocates were classified following four process types based on Halliday and Matthiessen (2014) – namely, material, mental, verbal, behavioural – each of which are defined below.

i. *Material processes* relate to physical action and have a material outcome. They can be either creative (i.e., brings about something new) or transformative (i.e., doing something to/changing something that already exists) processes.
ii. *Mental processes* refer to internal mental states and are grouped into four subcategories: perception (e.g. seeing, hearing), cognition (e.g. knowing), emotion, and desire/wanting.
iii. *Verbal processes* relate to communications more broadly. These can include verbs like ‘scream’ which indicates volume and lie which indicates something about the speaker’s intention.
iv. *Behavioural processes* relate specifically to physiological actions. These processes allow the distinction between mental processes (e.g. see) and the outward manifestation of these (e.g. watch). They also include physical actions for mental states (e.g. laugh, cry).

We used a collocation window of 4 words to the left and 4 words to the right of the node (e.g. the word that referred to social agents) and focused on the left collocates for adjectives and right collocates for verbs. We used the squared variant of the Mutual Information statistic (MI^2^) to determine the strength of a collocation, with a minimum score of 3 and a minimum frequency of 5, following (McEnery, 2004).

## 3. Results

### 3.1 Demographics and PSYRATS Scores

The mean age of participants was 46.0 years old. In terms of ethnicity of participants included in the study, the majority identified as being White British (N=11) followed by British African or Caribbean (N=3).The remaining participants were made up people who were White European (N=2), British Asian (N=2), British Latino (N=1), and mixed heritage (N=1). Of the total participants included in the analysis, 14 reported hearing hallucinated voices in the last two weeks allowing an assessment using the PSYRATS-V. The mean score was 19.4 with a mean length of time hearing voices of 17.4 years. Reports on the number of voices heard by participants ranged from 1 to 284 voices. Researchers identified 12 participants with delusions, with a mean PSYRATS-D score of 9.5. The mean length of time of the beliefs was 16.1 years.

### 3.2 Social agent characterisation

#### 3.2.1 Types of social agents

There were 1551 references to vocal social agents and 1365 references to non-vocal agents. For both types of agents, third person singular (‘he’, ‘she’, ‘it’) and plural pronouns (they, them) were the most commonly used in the corpus. The noun ‘voice(s)’ was the second most common reference (N = 236) to vocal social agents. The pronoun ‘they’ and the noun ‘voices’ were also present in the keyword list indicating that these words were significantly overused in the clinical corpus compared to the reference corpus. Both types of agent were frequently referred to by the noun ‘people’ (vocal social agent N = 93; non-vocal social agent N = 81). The 10 most frequent labels associated with both tags are listed in Table 1.

**Table 1.** Most frequent words used to reference illusory social agents in the interview texts

| *Type* | *Frequency referring to*<br>*Vocal Social Agents* | *Frequency referring to*<br>*Non-Vocal Social Agent* |
| --- | --- | --- |
| they | 404 | 291 |
| voice(s) | 236 | - |
| he | 192 | 128 |
| it | 75 | 139 |
| them | 125 | 89 |
| she | 76 | 117 |
| people | 93 | 81 |
| him | 47 | 33 |
| her | 26 | 41 |
| one(s) | 32 | 25 |

Overall, the pronoun ‘they’ was the most frequent in both tags with a count of 404 for vocal social agents and 291 associated with non-vocal social agents. This pronoun was used in reference to human and supernatural illusory social agents which were the most common type of social agents in participants’ experiences. Perdue et al. (1990) suggest that plural pronouns such as ‘we’, ‘us’, ‘they’, ‘them’ are linked to categorization of people or agents as part of ingroup or outgroup. Third person plural pronouns such as ‘they’ and ‘them’ are references for outgroup members. In the corpus, the ingroup designator ‘we’ was only used a total of 30 times and in all cases, except one, were in the context of the illusory social agents’ direct speech referencing their own collective (e.g. “we will kill you”, “we know what you’ve done”). Only one participant used the pronoun ‘we’ in reference to the activities of an army that she felt part of (“we’re involved in new world order”).

Darics and Koller (2019) highlight that ‘elite’ social actors are likely to be represented as individuals whereas those who are less prominent or ‘ordinary’ tend to be assimilated into collectives. Among participants whose experiences involved numerous social agents, only social agents who were experienced positively by the participants, or those that were perceived to be particularly powerful, frightening or to have higher levels of intelligence were referred to with singular pronouns. References to social agents as a group can be seen in references such as ‘voices’, ‘people’, ‘they’, ‘them’. The male third person singular pronoun ‘he’ was more frequent in both vocal social agents and non-vocal social agents than the female equivalent ‘she’. This finding is line with the results of previous voice hearing studies which found that although participants report hearing male, female and children’s voices, the identity of voices was frequently described as male (Corstens and Longden, 2013; McCarthy-Jones et al., 2014).

#### 3.3.1 Identities of agents

##### 3.3.1.1 Vocal agents’ identities

Due to the high number of pronominal references and the difficulty of differentiating each specific referent, only a broad estimate of the range of illusory social agents featured in the corpus can be obtained. Some participants were living with several types of illusory social agents (e.g. ghosts, people, animals) and tended to use plural pronominal forms and third person pronouns when describing these. The broad estimates discussed below were based on participants use of nominal forms (e.g. proper names, terms such as ‘spirits’, ghosts’, ‘man’) to label social agents.

In the clinical corpus, the most common type of illusory vocal social agents experienced aurally were human, with approximately 40 different labels (e.g. “person”, “guy”) used to describe them. The majority of these were internally individuated agents (Wilkinson and Bell, 2016) recognized by individual characteristics such as physical traits, gender, or race (e.g. “guy very short”, “white girl”) but without a named identity that would be recognised by others. Only two participants used first names to identify their vocal social agents. In both cases, although the participants heard multiple voices, only the social agents who were experienced as supportive were named. There are also several instances in the clinical corpus where social agents are aggregated into groups or referred to by a collective and represented without individuation (e.g. “two others, “people”, “bunch of guys”) (Wilkinson and Bell, 2016).

Nearly half of the human vocal social agents were externally individuated, i.e. they were associated with identities from the wider social world (Wilkinson and Bell, 2016). These included relatives, neighbours, former acquaintances, and service people from places such as cafes and public services frequented by the participant. Externally individuated illusory social agents were often referred to by their relation to another social actor (e.g. “dad”, “sister in law”, “wife”). In a minority of instances social agents were also identified by their social function or role (e.g. “waitress”, “umpire”).

Vocal social agents that were supernatural or animals were reported using just 8 and 4 labels, respectively. Supernatural beings were variously described by participants as “demons”, “spirits” and “ghosts”. Others were less clear on what the social agents were but felt that they were more than human. These social agents were simply referred to as “things”, “something” of a “demonic nature”, or part of a “higher power”. Only one participant reported illusory social agents that were animals that spoke to him. There were a minority of instances, captured by 4 reference types (e.g. refrigerator), where participants reported hearing sounds from inanimate objects.

##### 3.3.1.2 Non-vocal agents’ identities

This encompassed experiences of illusory social agents in delusions as well as visual and tactile hallucinations. There were 1365 references to these agents in the clinical corpus. As in the previous section, broad estimates of the types of social agents were obtained on the basis of nominal forms used to label them.

Similarly to the reports of vocal agents, the majority of these social agents were described as human using approximately 45 different reference types. Of these, over half were externally individuated agents. These agents included relatives, former acquaintances, members of a former cult, and ex-partners. These agents were more often referred to by their personal names and by their relation to another social agent or the participant (e.g. “mother-in-law”, “daughter”, “neighbour”). In several instances, illusory social agents were identified based on their social function or role (e.g. “doctors”, “prison guards”, “telepaths”, “umpires”) and specific traits, such as gender, stature (e.g. “little boy”; “girl”, “black woman”). There were two instances where internally individuated agents were referred to as a collective (e.g. “my army”).

Illusory social agents described as human but unknown to the participant were typically talked about as a collective (e.g. “people”, “people in black jackets”). Three participants identified social agents by their first or full names (e.g. “Jimi Hendrix”, “Robbie”). Specific names were only used in cases where the social agents were celebrities. In a minority of cases, social agents were identified by their physical traits such as age, race, and gender (e.g. “white person”, “girl”). One participant identified the same social agent by their function (“IT guy”, “personal trainer”) and in terms of their relationship to another social agent (“lover”).

Supernatural beings were involved in a minority of non-aural experiences and described using 24 reference types (e.g. “skeleton”, “spirit”). These were all, with the exception of one, described as evil. In two instances, participants described knowing the supernatural being with one participant stating that the “devil” came to her in a family member’s form and another saying that the “evil spirit” was also part of himself. Illusory social agents that were labelled with animal names were in the minority and described using approximately 12 reference types. These included anthropomorphous animals (e.g. “monkey man”, “mice with glasses”) as well as general animals (e.g. birds, slugs). A minority of experiences, captured by 11 reference types, involved social agents grouped as ‘other’ which included references to cartoon characters and objects that moved (e.g. “popeye”, “dancing flashing lights”).

#### 3.3.2 Intentions, Behaviours and Activities of Agents

This section focuses on the verb collocates of illusory social agents in a window of 0 words to the left and 4 words to the right. These collocates reflect what illusory social agents are represented as doing, and the intentions ascribed to them by participants.

##### 3.3.2.1 Vocal Agents’ Intentions, Behaviours and Activities

Table 3 lists the activities vocal social agents were reported as doing, MI^2^ score for each ‘lemma’ collocate, meaning that a particular collocate was realised in the data with different inflections (e.g. the lemma collocate ‘say’ was realised in the clinical corpus as ‘said’, ‘say’, ‘says’, and ‘saying).

**Table 3.** Activities of Vocal Social Agents. Words in bold are those that were identified as significantly more frequently in comparison to the reference corpus.

| *Verb*<br>*Collocate* | *Value*<br>*(MI<sup>2</sup>)* | *Dispersion*<br>*in corpus* | *Context* |
| --- | --- | --- | --- |
| **Say** | 11.36 | 15 | "they say different things you know the voices" -P15 |
| **Tell** | 10.17 | 8 | "they're uh .. just...telling me how it is you know"-P4<br>"the voices told me that I have to leave because I'm not safe"<br>-P1 |
| **Talk** | 9.59 | 8 | "they talk amongst themselves if you like" -P10 |
| **Argue** | 9.121 | 3 | "group of people that lived next door that would argue about<br>me" -P9 |
| **Insult** | 8.95 | 1 | "people insulting or bullying me" -P9 |
| **Defend** | 8.92 | 1 | "he was defending me as a friend" -P9 |
| **Try** | 8.52 | 8 | "they try to make trouble for me" -P8 |
| **Don't** | 8.18 | 5 | "they don't let me out" -P2<br>"they don't really care" -P9 |
| **Go** | 8.18 | 9 | "they go away sometimes" -P2<br>"he goes through the bible revelations with me" -P8 |
| **Know** | 7.85 | 4 | "they know I'm talking about them" P2 |
| **Shout** | 7.79 | 3 | "they're shouting"-P5 |
| **Get** | 7.56 | 6 | "We'll get you out of your flat if it's the last thing we do" -P3 |
| **Call** | 7.53 | 2 | "she calls me names"-P8 |
| **Bring** | 7.51 | 1 | "they bring me down sometimes"- P17 |
| **Can** | 7.24 | 7 | "he could be American using a London accent" -P18 |
| **Want** | 7.21 | 6 | "they want to protect me from bad people" -P1 |
| **Do** | 7.19 | 9 | "he can do my head in" -P8 |
| **Keep** | 7.04 | 2 | "he keeps disturbing my life" -P17 |
| **Make** | 6.95 | 8 | "voices make me out to be inadequate" -P3<br>"they try and make me cry" -P4 |
| **Come** | 6.94 | 6 | "she'll come and help me" -P8 |
| **Start** | 6.88 | 5 | "they start on me even more" -P15 |
| **Have** | 6.69 | 8 | "the voices have colour to them" -P4 |
| **Speak** | 6.62 | 4 | "people speak to me in the street" -P3 |
| **Pull** | 6.21 | 2 | "they will pull you" -P5 |
| **Sit** | 6.18 | 2 | "he's sat there listening now" -P18 |
| **Listen** | 6.17 | 3 | "they say they can listen" -P8 |
| **Kill** | 6.15 | 5 | "he said kill him kill him"- P7 |
| **Laugh** | 5.98 | 4 | "he is laughing through me" -P18 |
| **Leave** | 5.98 | 2 | "he won't leave me alone" -P17 |
| **Think** | 5.91 | 5 | "he goes pahaha at whatever he thinks is funny"- P18 |
| **Give** | 5.74 | 4 | "they give me bad thoughts"- P2 |
| **Bully** | 5.73 | 1 | "they were bullying me about rape" -P9 |
| **Help** | 5.55 | 5 | "his voice will help me" -P14 |
| **See** | 5.02 | 3 | "people can see me watching it" -P9 |
| **Take** | 4.09 | 5 | "the voices take me through the forest" -P1 |

Predictably, nearly half the verb collocates associated with vocal social agents were verbal processes that took place either between social agents or social agents and participants. Seven collocates (‘say’, ‘tell’, ‘talk’, ‘explain’, ‘speak’, ‘ask’, ‘take’) represented illusory social agents as engaging in interactive conversations either with other agents or with the participants. The collocate ‘take’ was counted both as a verbal process, because in context it is a colloquial metaphorical expression denoting the voices verbally guiding the participant through something, and as a material process in accordance with the basic meaning of ‘take’. Here, basic meaning refers to a current meaning listed in the dictionary that is either more concrete (what they evoke is easier to imagine, see, hear, feel, smell, and taste), related to bodily action, or more precise, as opposed to vague (Pragglejaz Group, 2007).

Eight collocates capture verbal processes that showed illusory social agents in communicative acts that could be described as negative or unpleasant for the participants: ‘argue’, ‘shout’, ‘insult’, ‘bully’, ‘call’ (name calling), ‘go’, ‘start’ and ‘make’. Some of these, such as insulting and bullying (by means of name-calling and spreading rumours), were particularly prominent for one participant (P9) whose experience was dominated by public shaming. The collocate ‘make’ (both a verbal and material process) showed illusory social agents belittling participants (e.g. “they make rude comments about me” -P3). ‘Start’ was used in a colloquial way to indicate the beginning or intensifying of verbal harassment from social agents (e.g. “they start on me even more” -P15), while ‘goes’ was used similarly to describe the social agent narrating, and possibly discussing, passages from a bible chapter which the participant disliked because of the apocalyptic themes within.

A number of collocates represented material processes that vocal social agents performed. The collocates ‘do’, ‘keep’, ‘pull’, ‘bring’, ‘go’, ‘leave’, ‘come’, ‘make’, ‘help’ are transformative material processes that portrayed illusory social agents as influencing participants’ mental and physical wellbeing and environment. Eight of these actions implicitly characterised the agents as intrusive and unwelcome characters that negatively impacted on participants mental wellbeing (e.g. “they *bring* me down” -P4). Three participants reported social agents being able to physically touch them by pulling on or attacking their bodies. In addition, the negated auxiliary ‘don’t’ was frequently used to describe social agents restricting participants’ movements (e.g. “they *don’t* let me out” -P2). Illusory social agents were also represented as independent beings that were able to come and go from participants’ physical and mental spaces of their own volition. The collocate ‘help’ portrayed social agents as offering support and comfort to participants. One collocate (‘give’) was a creative material process because illusory social agents were represented as bringing about something to the participants cognition (e.g. “they *give* me bad thoughts” -P2).

Two collocates (‘laugh’, ‘listen’) highlighted behavioural processes of illusory social agents. As described by Thompson (2013), behavioural processes capture outward signs of mental processes and mental states. These collocates are the communicative or interactive result of illusory social agents’ mental processes such as being able to hear, experience different mental states and humour.

Five collocates (‘see’, ‘know’, ‘think’, ‘want’, and ‘try’) captured mental processes of illusory social agents. In terms of perception, participants reported social agents being able to see participants’ activities and bodies. Participants experienced this as intrusive because it was against their wishes and interfered with their ability to engage in certain activities (e.g. being intimate with partners). Social agents were characterised as knowing and thinking beings that were aware of the participants history (e.g. “they *know* already about me… that I’m bad” -P1) and present activities (e.g. “they *know* I am talking about them” -P1). One participant described her voice as having plans (e.g. “he *thinks* he’s going to get the money back” -P18) and being able to physically laugh through the participant at things the social agent thought funny.

Finally, some participants made inferences about the intentions and/or desires of the social agents using verbs such as ‘try’ and ‘want’.

##### 3.3.2.2 Non-Vocal Agents’ Intentions, Behaviours and Activities

Verb collocates of non-vocal illusory social agents are shown in Table 4. The list of collocates suggests that these agents were engaged in more material and behavioural processes than the vocal social agents above. They were also involved in verbal processes, however, to a much lesser degree than vocal agents: only four verb collocates (‘talk’, ‘say’, ‘speak’, and ‘tell’) were verbal processes. With these agents, communications directed at participants did not always come directly in spoken form. Rather they were often transmitted through various modes (e.g. television, lights, intuition). Some participants did not hear agents communicating but felt certain that social agents were talking about them while others could hear social agents mocking or making plans to harm them. One participant communicated with illusory social agents through online chat forums.

**Table 4.**
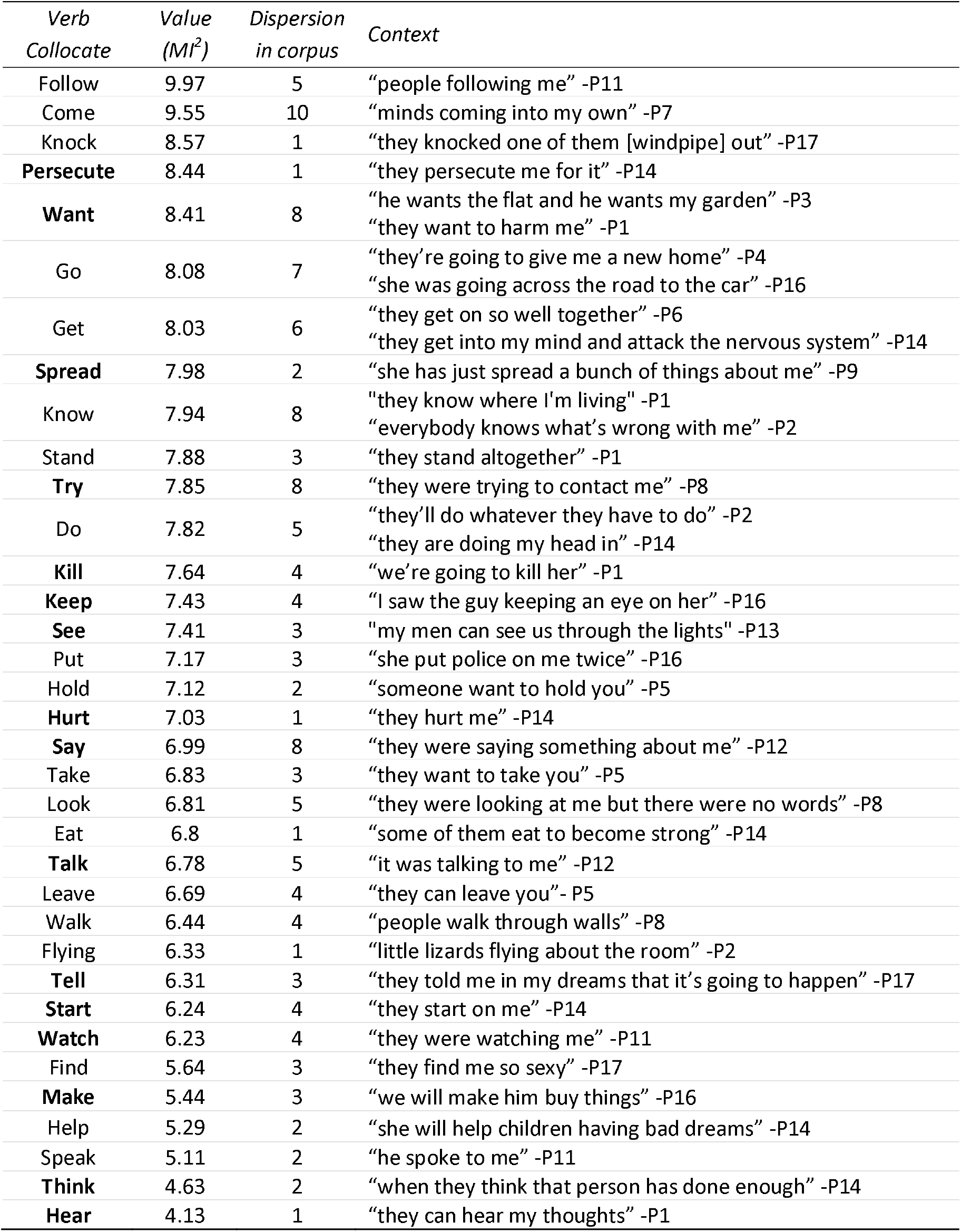
Activities of Non-Vocal Social Agents. Words in bold are those that were identified as significantly more frequently in comparison to the reference corpus.

| *Verb Collocate* | *Value (MI<sup>2</sup>)* | *Dispersion in corpus* | *Context* |
| --- | --- | --- | --- |
| Follow | 9.97 | 5 | "people following me" -P11 |
| Come | 9.55 | 10 | "minds coming into my own" -P7 |
| Knock | 8.57 | 1 | "they knocked one of them [windpipe] out" -P17 |
| **Persecute** | 8.44 | 1 | "they persecute me for it" -P14 |
| **Want** | 8.41 | 8 | "he wants the flat and he wants my garden" -P3<br>"they want to harm me" -P1 |
| Go | 8.08 | 7 | "they're going to give me a new home" -P4<br>"she was going across the road to the car" -P16 |
| Get | 8.03 | 6 | "they get on so well together" -P6<br>"they get into my mind and attack the nervous system" -P14 |
| **Spread** | 7.98 | 2 | "she has just spread a bunch of things about me" -P9 |
| Know | 7.94 | 8 | "they know where I'm living" -P1<br>"everybody knows what's wrong with me" -P2 |
| Stand | 7.88 | 3 | "they stand altogether" -P1 |
| **Try** | 7.85 | 8 | "they were trying to contact me" -P8 |
| Do | 7.82 | 5 | "they'll do whatever they have to do" -P2<br>"they are doing my head in" -P14 |
| **Kill** | 7.64 | 4 | "we're going to kill her" -P1 |
| **Keep** | 7.43 | 4 | "I saw the guy keeping an eye on her" -P16 |
| **See** | 7.41 | 3 | "my men can see us through the lights" -P13 |
| Put | 7.17 | 3 | "she put police on me twice" -P16 |
| Hold | 7.12 | 2 | "someone want to hold you" -P5 |
| **Hurt** | 7.03 | 1 | "they hurt me" -P14 |
| **Say** | 6.99 | 8 | "they were saying something about me" -P12 |
| Take | 6.83 | 3 | "they want to take you" -P5 |
| Look | 6.81 | 5 | "they were looking at me but there were no words" -P8 |
| Eat | 6.8 | 1 | "some of them eat to become strong" -P14 |
| **Talk** | 6.78 | 5 | "it was talking to me" -P12 |
| Leave | 6.69 | 4 | "they can leave you" -P5 |
| Walk | 6.44 | 4 | "people walk through walls" -P8 |
| Flying | 6.33 | 1 | "little lizards flying about the room" -P2 |
| **Tell** | 6.31 | 3 | "they told me in my dreams that it's going to happen" -P17 |
| **Start** | 6.24 | 4 | "they start on me" -P14 |
| **Watch** | 6.23 | 4 | "they were watching me" -P11 |
| Find | 5.64 | 3 | "they find me so sexy" -P17 |
| **Make** | 5.44 | 3 | "we will make him buy things" -P16 |
| Help | 5.29 | 2 | "she will help children having bad dreams" -P14 |
| Speak | 5.11 | 2 | "he spoke to me" -P11 |
| **Think** | 4.63 | 2 | "when they think that person has done enough" -P14 |
| **Hear** | 4.13 | 1 | "they can hear my thoughts" -P1 |

Twenty-two collocates associated with these agents are material processes where a social agent was doing something to a participant or another social agent. Most of these collocates were transformative material processes as they represented illusory social agents affecting participants’ bodies and environments. Nine of these (‘hurt’, ‘persecute’, ‘start’, ‘knock’, ‘hold’, ‘do’, ‘stand’, ‘get’, and ‘kill’) represented the agents as causing physical hurt and mental distress to participants or restricting their movements (as in illusory social agents standing in someone’s way, preventing their access to their kitchen and opening the front door). The collocate ‘get’, when used as a material process, portrays social agents entering the minds of participants and attacking them from within.

Six collocates of transformative processes (‘come’, ‘go’, ‘leave’, ‘fly’, ‘walk’, ‘follow’) depicted social agents as moving around space. Similar to the vocal agents in the previous section, non-vocal agents were portrayed as independent and mobile beings who were able to freely enter and leave physical and mental spaces regardless of participants wishes. Two collocates (‘make’, and ‘help’) illustrated social agents’ attempts to influence the actions/situations of real and illusory agents. For example, ‘help’ was used to characterise an illusory social agent’s abilities to support people in need. Three collocates ‘spread’, ‘take’, ‘put’) were creative material processes that brought about something (e.g. smells on bodies or environment) or someone (e.g. police) new. The collocate ‘take’ was used in the context of illusory social agents taking naked pictures of a participant and distributing it among other illusory social agents, but also denote social agents taking over participants’ brain or agency.

Two collocates highlighted behavioural processes of social agents. These verbs portrayed social agents observing participants (‘watch’, ‘look’). More mental processes were attributed to the non-vocal social agents when compared to vocal agents. In terms of perception, social agents were able to hear and see participants. Cognitively these agents were also portrayed as knowing and thinking beings who were aware of participants’ history and current activities. A minority of participants commented on the mental states of illusory social agents as captured by the collocates ‘get’ and ‘find’. For example, one participant felt her social agents were attracted to her (“they *find* me so sexy” -P17) and associated these feelings to be the driver behind the social agents constantly trying to touch her body. Another participant reported social agent got angry with him and a third commented on social agents’ affections for each other (“they *get* on so well together” -P6). Social agents were perceived to have motivations, plans and desires, as represented by the collocates ‘try’, ‘want’, ‘going’ to.

## 4. Discussion

This study aimed to characterise the experience of illusory social agents in psychosis by using computational corpus linguistics to identify the characteristics of agents as described in open-ended interviews. The results showed that social agents: (i) are represented with varying levels of richness in participants’ experiences, (ii) are attributed with different kinds of identities including physical characteristics and names, (iii) are perceived to have internal states and motivations that are different from those of the participants, and (iv) interact with participants in various ways including through communicative speech acts, affecting participants’ bodies and moving through space. Overall, illusory social agents were characterised in participants’ experiences as active and dynamic beings who engaged in a range of verbal, mental, behavioural and material processes across both vocal and non-vocal hallucinations and in delusions.

The results reported here largely support and extend Alderson-Day et al. (2020) findings on personification in hallucinated voices from patients in first-episode psychosis services. They reported that voice hearers ranged in their experiences that voices were largely without agency (i.e. they appear as unresponsive words or sounds) to those with complex personified identities associated with a capacity for interactive communication and companionship. Here, we report on the experience of both hallucinated voices and non-vocal illusory social agents in psychosis, and how they are experienced as ‘thinking, behaving and interacting’ as described by the participants.

Collocation analysis showed that illusory social agents were represented as omniscient, powerful, independent, and often malicious characters. Agents were also represented as having the power to influence and create changes to participants’ environment and wellbeing. Notably, it was not just what the agent did that mattered in terms of how they were experienced, but also what they did not do (e.g. “they don’t care”). The analysis also showed that that majority of the speech acts by voices were rapport-damaging acts designed to attack participants ‘face’ (Demjén et al., 2020) their sense of self-worth and their reputation. Illusory social agents in general also frequently infringed on participants’ sociality rights. Acts such as making threats, telling participants to harm others, warning participants that their environment is dangerous, and warning participants to not trust or communicate with others in their social world all interfere with participants’ right to associate with others and to be treated fairly.

In terms of the depth and complexity of agent representations, the majority were what (Wilkinson and Bell, 2016) described as ‘internally individuated’ – meaning they were identifiable by the individual based on the illusory social agents unique properties but did not correspond to any person or agent in the external world that others would recognise. However, illusory social agents were also endowed with motivations and mental states that were not fully accessible to the participants and to a level of complexity that is usually associated with human-level intelligence (Thompson, 2013). They were represented in the corpus as having an inner life consisting of thoughts, feelings, knowledge, intentions, and plans. Participants spoke of illusory social agents experiencing a range of feelings including anger, jealousy, sexual attraction, and pain/sadness, however, not all of these were captured in the collocates or keyword analysis. For the majority of the participants, particularly those on the wards, illusory social agents appeared frequently and were relatively dynamic characters that sometimes irritated or angered the participants but at other times offered comfort and humour. Based on the scalar model of minimal to complex personification of voices developed by Semino et al. (2020), these qualities of (i) having ‘online’ emotions, (ii) possessing internal states and motivations that are not accessible to the participants, (iii) engaging in interactions with participants, and (iv) having different behaviours, suggest that many social agents are personified in complex ways that are similar to the way real people are perceived in the shared external social world. Notably, Wilkinson and Bell (2016) suggested that ‘externally individuated’ illusory social agents in psychosis (i.e. those that do correspond to external identities) reflect a richer agent representation than internally individuated ones, but the evidence presented here suggests that internally individuated agents may be equally as rich in terms of their human-like agentive properties.

In a study which applied models of impoliteness to the experience of voice hearing, (Demjén et al., 2020) drew on linguistics concepts of ‘face’, sociality rights, and rapport enhancing / damaging acts (Spencer-Oatey, 2000) to illustrate how linguistic features associated with the exercise of power in relationships were present in how voices were described as attacking, maintaining or boosting voice hearers’ sense of self and relationships verbally. However, these processes were also evident when the activity of non-vocal agents was investigated. For example, several participants commented on the relentlessness of the social agents, feeling watched, being followed, prevention of going out, and touching body parts against participants’ consent. These acts interfered with participants’ ability and rights to create and maintain relationships with others in their real social world.

While the majority of work on social agents in psychosis has focused on hallucinated voices, following Bell et al. (2017), the results presented here suggests that agentive characteristics are similarly present in the delusions and non-auditory hallucinations of psychosis, potentially suggesting that altered social agent processing may be a frequent characteristic across delusions and hallucinations.

However, we also note that one characteristic used to describe voices was ‘strangeness’, something akin to perplexity (Humpston, 2014). For example, when asked about her voices one participant said “I don’t honestly know what the voices are” -P10. The presence of keywords such as “weird” (e.g. “it’s so weird and confusing to explain” -P12), “strange, and “random” used by participants trying to describe the experience may also be indicative of an ongoing processing of these agent-like experiences in light of the extent to which they do or do not align with existing schemas. This suggests that in many cases participants are using bottom up processes to integrate available information of social agents to build up agent representation.

## Data Availability

The #Lancsbox analysis files minus the original transcripts have been made available on the online archive: https://osf.io/4zwq8/

